# Cerebral Metabolic Rate of Oxygen and Accelerometry-Based Fatigability in Community-Dwelling Older Adults

**DOI:** 10.1101/2025.01.11.25320396

**Authors:** Emma L. Gay, Caterina Rosano, Paul M. Coen, Nicholaas Bohnen, Theodore Huppert, Yujia (Susanna) Qiao, Nancy W. Glynn

## Abstract

Alterations in energy metabolism may drive fatigue in older age, but prior research primarily focused on skeletal muscle energetics without assessing other systems, and utilized self-reported measures of fatigue. We tested the association between energy metabolism in the brain and an objective measure of fatigability in the Study of Muscle, Mobility and Aging (N=119, age 76.8±4.0 years, 59.7% women). Total brain cerebral metabolic rate of oxygen (CMRO_2_) was measured using arterial spin labeling and T_2_-relaxation under spin tagging MRI protocols. Accelerometry-based fatigability status during a fast-paced 400m walk was determined using the Pittsburgh Fatigability Index (PPFI, higher=worse). Confounders included skeletal muscle energetics, measured in vivo using spectroscopy and ex vivo using respirometry, cardiorespiratory fitness (VO_2_peak), weight, medication count, and multimorbidity.

Multivariable logistic regression models were used to estimate the association (odds ratio (OR)) of CMRO_2_ with PPFI>0 compared to the referent group PPFI=0. Models were first adjusted for age and sex, and further adjusted for confounders. In this sample, 41.2% had PPFI>0 (median 3.3% [0.4-8.0%]). CMRO_2_ was positively associated with PPFI>0 (age and sex adjusted OR=1.61, 95% CI: 1.06, 2.45, p=0.03); adjustment for confounders attenuated the association. The positive association of brain energetics and fatigability warrants further study in older adults.

## Introduction

Fatigability, the degree one is limited by fatigue during a walking task, is an established measure of impairment in older adults (Enoka et al. 2021; Van Geel et al. 2020) and a subclinical indicator of functional limitations (Schnelle et al. 2012; Hunter 2018; Qiao et al. 2023). We have recently reported that both lower skeletal muscle energetics and cardiorespiratory fitness (CRF, VO_2_peak) were associated with greater accelerometry-based fatigability in older adults (Qiao et al. 2023; Qiao, Santanasto et al. 2023). Although the central nervous system plays an important role in the perception of fatigue (Stults Kolehmainen et al. 2020; Marcora 2019; Taylor et al. 2016), studies have primarily assessed neurological patients (Camandola & Mattson 2017; Dalsgaard & Secher 2007; Brooks & Martin 2014; Peralta et al. 2019; Zhang et al. 2018; West et al. 2020) with few reports in older adults without neurological diagnoses (Kato et al. 1999). Studying brain energetics in relation to performance fatigability may help understand the processes underlying this devastating and common phenomenon in older age.

Among the neuroimaging methods to capture brain energetics, T_2_-relaxation under spin tagging (TRUST) and Arterial Spin Labeling are emerging as non-invasive approaches to quantify Cerebral Metabolic rate of Oxygen (CMRO_2_) in a relatively short period of time, with demonstrated validity and reliability, and without contrast agents or radioactive labels (Alsop et al. 2015; Jiang et al. 2021; Xu et al. 2009; Vestergaard et al. 2017). CMRO_2_ reflects the amount of oxygen extracted by the brain parenchyma, and increases with age among adults without clinically overt diseases (Xu et al. 2009). Greater CMRO_2_ may indicate greater metabolic costs to maintain homeostasis, perhaps due to reduced cellular efficiency and/or in response to age-related impairments in other systems (Peng et al. 2014; Lu et al. 2011).

We examined the relation between CMRO_2_ and performance fatigability using the Pittsburgh Fatigability Index (PPFI) (Qiao et al. 2022). We hypothesized that those with higher CMRO_2_ would have greater PPFI. Since muscle energetics and CRF play a critical role in driving fatigability, we assessed to what extent these measures modified the association of CMRO_2_ with fatigability.

## Methods

### Study Sample

Older adults age ≥70 years enrolled in the Study of Muscle, Mobility and Aging (SOMMA, http://sommaonline.ucsf.edu) from the Pittsburgh clinical site (N=439) were recruited for the SOMMA-Brain Ancillary study. Exclusion criteria for the parent study included: inability to walk one-quarter of a mile or climb a flight of stairs; body mass index (BMI) ≥ 40 kg/m^2^; active malignancy or dementia; or medical contraindication to biopsy or magnetic resonance imaging (MRI). Additionally, participants had to be able to complete a usual-paced 400m walk (Cummings et al. 2023). To be eligible for the SOMMA-Brain Ancillary study the skeletal muscle biopsy had to have occurred within the past 12 months (n=285), and without diagnosed neurologic disorder. A total of 150 individuals agreed to participate in the ancillary study and completed the neuroimaging protocol (Figure 1) (Rosano et al. 2024). Of those, 26 did not complete the walking test needed to derive PPFI (19 were not tested due to scheduling issues, 2 could not be reached, 2 unable to finish walk, 1 technical error, 1 was ineligible, and 1 refused). Of the 124, 5 participants did not have useable accelerometry data for deriving the performance fatigability outcome. Thus, the final analytic sample was n=119 (Figure 1). The average time between muscle biopsy and neuroimaging was 9.4 months. The WIRB-Copernicus Group Institutional Review Board (IRB# 20180764) and the University of Pittsburgh Human Research Protection Office (PittPRO# 20110230) approved the study and all participants gave informed written consent.

**Figure 1:**
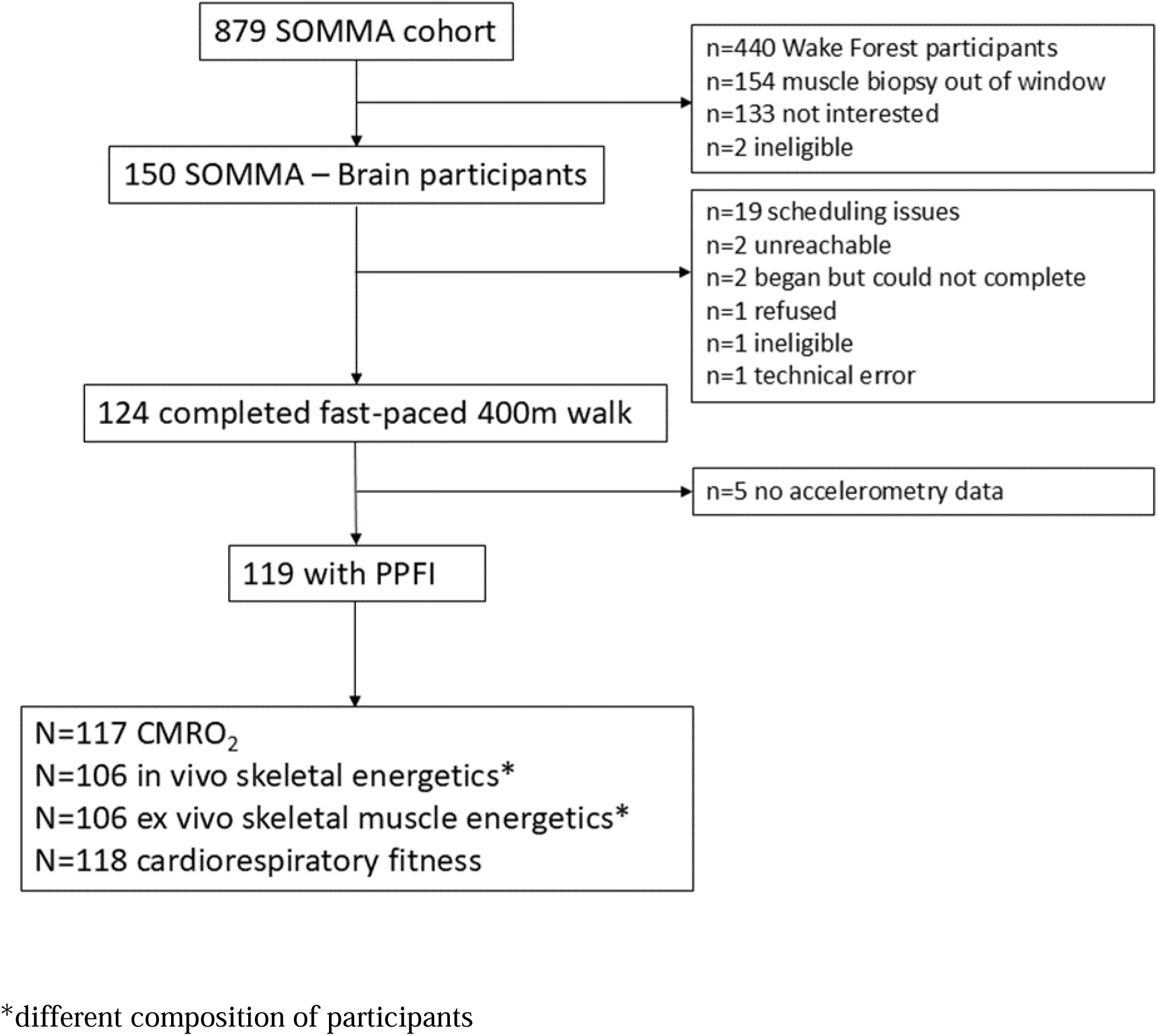
Flowchart for Inclusion and Exclusion of Participants from the Study of Muscle, Mobility and Aging (SOMMA) – Brain Ancillary Study in this Analysis

### Performance Fatigability

Participants wore an ActiGraph GT9X accelerometer (ActiGraph LLC) on both ankles during the fast-paced 400m walk. Triaxial raw accelerometer data were collected at a sampling frequency of 100Hz. During the fast-paced 400m walk, participants were instructed to walk as quickly as possible, without running, at a pace they could maintain for 10 laps on a 20m course (Simonsick et al. 2006).

Raw accelerometer data from the non-dominant ankle were processed in R to calculate PPFI (Qiao et al. 2022), a ratio comparing the area under the individual’s observed cadence-versus-time trajectory during the walk to a hypothetical area that would be observed if the individual’s maximum cadence were maintained throughout the walk. Individual-level smoothed cadence trajectories were fit using penalized regression splines. Specific details about the derivation of PPFI have been published (Qiao et al. 2022). Higher PPFI score (range 0-100%) indicates greater performance fatigability (Qiao et al. 2022). Participants who completed the fast-paced 400m walking within 5 minutes exhibited no performance fatigability during the walking task and thus, were classified as PPFI=0 (Qiao et al. 2022). PPFI was initially validated at the non-dominant wrist, but the non-dominant ankle was an appropriate substitution as identification of physical activity is 95% accurate for ankle worn devices (Mannini et al. 2013). Additionally, stride segmentation (i.e., cadence), which is an essential input to derive PPFI, is highly accurate for ankle worn accelerometry when using the ADEPT R package with estimated deviations for stride of 1.24% at the left ankle and 1.30% at the right ankle (Karas et al. 2021).

### Brain MRI - Cerebral Metabolic Rate of Oxygen (CMRO_2_)

CMRO_2_, the amount of oxygen consumed per unit mass and per unit time, depends on cerebral blood flow (CBF) and oxygen extraction fraction (OEF)(Xu et al. 2009). The Fick principle of arteriovenous oxygen difference provides an absolute measure of CMRO_2_ in the whole brain (Kety & Schmidt 1945; Lee et al. 2013). OEF, reflecting the proportion of O_2_ extracted from the blood, was estimated non-invasively via T_2_-relaxation under spin tagging MRI in the sagittal sinus and arterial oxygen saturation via pulse oximetry (Jiang et al. 2021). CBF, reflecting the supply of O_2_ to the brain, was assessed using arterial spin labeling perfusion MRI (Siemens Biograph mMR PET/MR)(Alsop et al. 2015).

### Cortical volume and white matter hyperintensity quantitation

MRI volumes were distortion-corrected, registered, and segmented as described in (Glasser et al. 2013) using a combination of FSL and FreeSurfer analysis programs. Total and subcortical gray matter volumes were obtained via FreeSurfer (Dale et al. 1999). Regions were labeled with reference to the Desikan atlas (Desikan et al. 2006). Gray matter atrophy was calculated as total gray matter volume/intracranial volume and subcortical gray matter atrophy as subcortical gray matter volume/intracranial volume. White matter hyperintensity (WMH) on MRI, a measure of cSVD (cerebral small vessel disease), was quantitated via automated segmentation methods as previously described (Schmidt & Wink 2017).

### Skeletal Muscle Energetics

#### Maximal ATP Production

Based on Qiao et al. finding that lower maximal ATP production (ATPmax) was associated with higher PPFI during a usual-paced 400m walk in the SOMMA parent study (Qiao, Santanasto, et al. 2023), we examined this measure as a potential covariate in our models. ATPmax was quantified using ^31^P magnetic resonance spectroscopy to measure the rate of phosphocreatine (PCr) regeneration following a short bout of exercise. A 3 Tesla MRI scanner (Siemens Medical System – Prisma) using a 12” dual-tuned, surface radiofrequency coil (PulseTeq, Limited) placed over the right distal vastus lateralis was used to collect ^31^P spectra. Participants performed two bouts of isometric knee extension against the resistance of an ankle strap as previously described (Cummings et al. 2023). PCr recovery rate after exercise was fit and the time-constant of the mono-exponential fit (tau) was used to calculate ATPmax (Blei et al. 1993; Jubrias et al. 2003; Amara et al. 2008). In SOMMA, the mean coefficient of variation for duplicate measures of ATPmax was 9.9% across clinic sites (Mau et al. 2023).

#### Skeletal Muscle Respiration

Our previous work in SOMMA also revealed that lower maximal complex I & II supported oxidative phosphorylation (max OXPHOS) and maximal electron transport system (max ETS) were associated with higher PPFI during a usual-paced 400m walk (Qiao, Santanasto, et al. 2023), thus we also evaluated these two measures of skeletal muscle respiration as potential covariates. A skeletal muscle biopsy was taken from the medial vastus lateralis after a 12-hour fast and limited exercise for 48 hours prior to the procedure (Zamora et al. 2024). Approximately 20mg of the specimen was placed in a biopsy preserving solution for high-resolution respirometry (Zamora et al. 2024). Approximately 2-3 mg of myofiber bundles were then weighed and placed into Oxygraph-2K respirometer chambers (O2K, Oroboros Instruments, Austria). Assays were run in duplicate at 37°C within a specific range of O_2_ concentrations (400-200 μm). Steady-state oxygen flux was normalized to the fiber bundle wet weight using Datlab 7.4 software (Coen et al. 2013; Mau et al. 2023). Technician was controlled for in analysis.

### Cardiorespiratory Fitness

Cardiorespiratory fitness was measured by cardiopulmonary exercise testing (CPET) using a modified symptom-limited Balke treadmill protocol where speed and grade increased incrementally (Wolf et al. 2024). After a 5-minute preferred walking speed treadmill task, testing for VO_2_peak began with incremental rate (0.5 mph) and/or slope (2.5%) increases in 2 minute stages until respiratory exchange ratio was ≥ 1.05 and Borg Rating of Perceived Exertion was ≥17. Absolute VO_2_peak was determined in the BREEZESUITE software as the highest 30-second average of VO_2_ (mL/min) achieved (Wolf et al. 2024). Both absolute and weight adjusted (mL/kg/min) VO_2_peak were used in analyses as appropriate.

### Covariates

Age in years, brain atrophy, white matter hyperintensities and joint pain in the last month were measured during the baseline MRI visit of SOMMA-Brain as previously described (Rosano et al. 2024). Measures collected during the SOMMA baseline visit of the parent study included sex and weight measured using a balance beam or digital scale, without shoes and with light clothing. Participants reported the prescription medications they had taken in the past 30 days, a count of medications was used in this analysis. Self-reported history of physician diagnosed chronic health conditions and depressive symptoms were combined to create the SOMMA multimorbidity index, which was dichotomized to 0-1 and >1 for this analysis.

### Statistical Analysis

Characteristics of the participants by performance fatigability status (PPFI=0 vs PPFI>0) were compared using ANCOVA (continuous) or logistic regression (categorical); reported p-values reflect age and sex adjustment. We examined associations between CMRO_2_, PPFI (continuous), and variables of interest that were significantly different by performance fatigability status using partial Pearson and Spearman (PPFI) correlations adjusted for age and sex. Logistic regression was used to examine the association between CMRO_2_ and performance fatigability status adjusted for age, sex, plus skeletal muscle energetics (and technician for ex vivo measures) in separate models. Next, we adjusted for cardiorespiratory fitness, weight; and last, for any other variables that were significantly different by performance fatigability status. The units of the explanatory variable, CMRO_2_, were scaled to one standard deviation (SD) for interpretation, Additionally, ATPmax, max OXPHOS, max ETS and VO2peak were entered into models as standardized variables. Analyses were conducted in SAS version 9.4 using the May 2024 SOMMA data release.

## Results

We recruited 119 adults (age 76.8±4.0 years, 59.7% women) from the Study of Muscle, Mobility and Aging (Cummings et al. 2023) with data on CMRO_2_, fatigability, muscle energetics and CRF. Median PPFI score was 1.3% (range 0-8.0%) in the full sample, and 41.2% (n=49) had PPFI>0, median of 3.3% (range 0.4-8.0%) (Table 1). Compared to those with PPFI=0 (n=70), those who exhibited fatigability, were 2.6 years older and 16.5% more likely to be a woman. Those with PPFI>0 had 7.9% higher CMRO_2_, 5.9 mL/kg/min lower VO_2_peak, 7.5 kg higher weight, reported taking 1.5 more medications and were 16.4% more likely to have multimorbidity >1 compared to those with no fatigability, all p<0.05 (Table 1). Muscle energetics (in vivo and ex vivo), brain atrophy, normalized white matter hyperintensities, and presence of joint pain were similar by fatigability status, p>0.05 (Table 1).

**Table 1:** Characteristics of Participants by Fatigability Status: Study of Muscle, Mobility and Aging (SOMMA) – Brain Ancillary Study.

| **Variable** | **No Performance**<br>**Fatigability**<br>**(PPFI=0)**<br>**n=70** | **Exhibiting**<br>**Performance**<br>**Fatigability**<br>**(PPFI>0)**<br>**n=49** | **p-value\*** |
| --- | --- | --- | --- |
| Age, years | 75.7 ± 2.9 | 78.3 ± 4.7 |  |
| Sex, Women | 37 (52.9) | 34 (69.4) |  |
| Cerebral Metabolic Rate of Oxygen (CMRO<sub>2</sub>), µmol/100 g/min | 10.5 ± 2.8 | 11.4 ± 2.9 | 0.03 |
| ATPmax, mM/sec | 0.60 ± 0.20 | 0.53 ± 0.10 | 0.06 |
| Maximal Complex I&II supported OXPHOS, pmol/(s*mg) | 71.4 ± 22.7 | 63.5 ± 16.1 | 0.1 |
| Maximal Electron Transport System Capacity, pmol/(s*mg) | 86.2 ± 25.5 | 76.8 ± 18.9 | 0.09 |
| VO<sub>2</sub>peak, mL/kg/min | 25.6 ± 5.4 | 19.7 ± 3.8 | <0.0001 |
| Weight, kg | 69.6 ± 12.7 | 77.1 ± 14.3 | <0.0001 |
| Medication count | 4.0 ± 3.3 | 5.5 ± 3.9 | 0.02 |
| Multimorbidity >1 | 6 (8.6) | 12 (25.0) | 0.008 |
| Brain atrophy (total gray/intracranial volume) | 0.33 ± 0.02 | 0.33 ± 0.03 | 0.9 |
| White Matter Hyperintensities** | 0.014 ± 0.02 | 0.018 ± 0.02 | 0.6 |
| Joint pain (yes/no) | 24 (34.3) | 15 (30.6) | 0.5 |

Values are mean ± standard deviation or n (%)
*Abbreviations:* ATPmax – maximal ATP production, OXPHOS – oxidative phosphorylation
\*Age and sex adjusted p-value from ANCOVA (continuous) or logistic regression (categorical)
\*\* normalized for total white matter

In age and sex-adjusted models, higher CMRO_2_ was correlated with higher PPFI score (r=0.26, p=0.01), lower VO_2_peak (r=-0.22, p=0.04), and multimorbidity >1 (r=0.35, p=0.0008), but not with muscle energetics, weight or medication counts (Figure 2).

**Figure 2:**
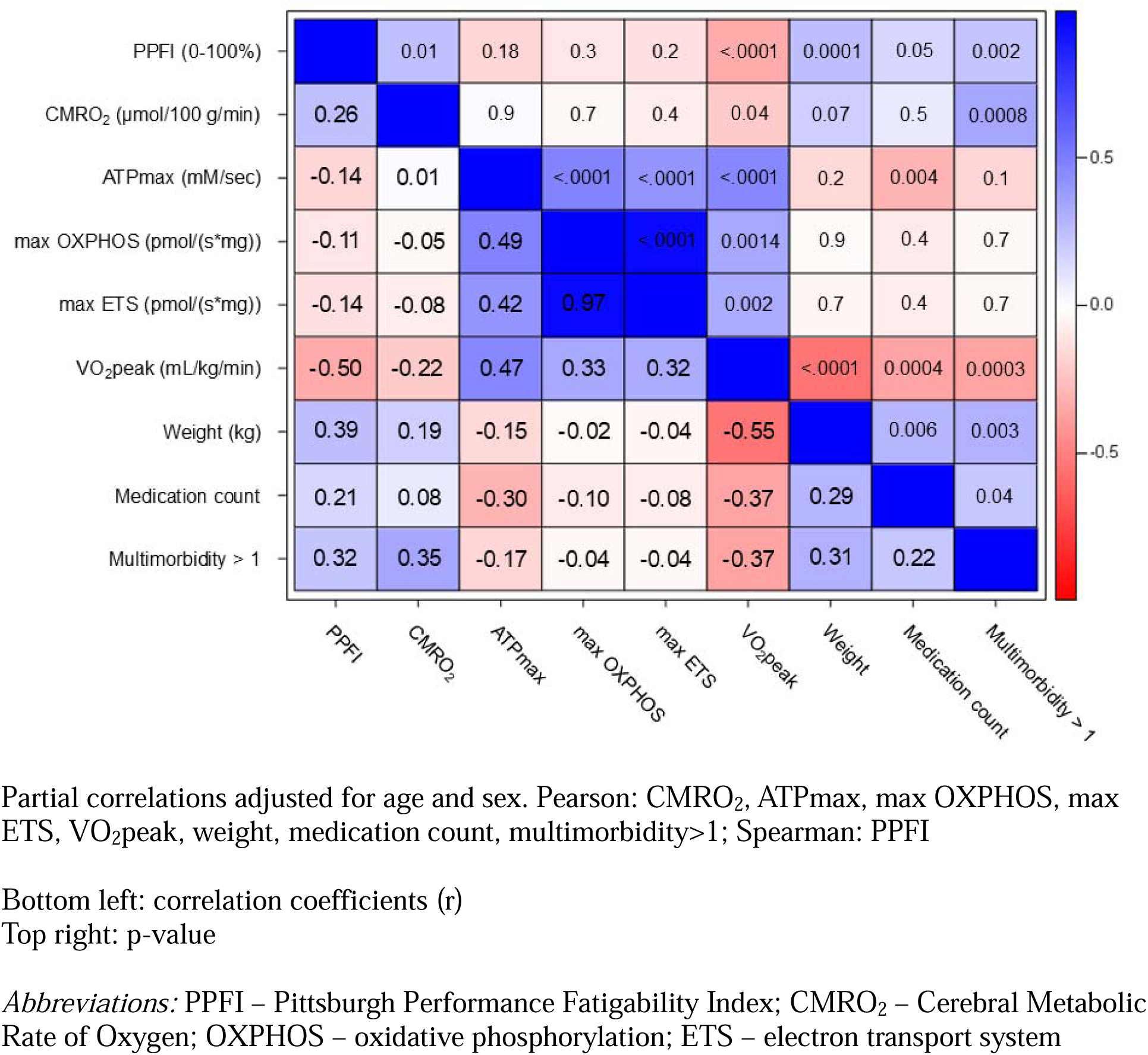
Partial Pearson and Spearman Correlations for the Pittsburgh Fatigability Index (PPFI) and Main Contributors of PPFI

In multivariable models, one SD higher CMRO_2_ (2.87 μmol/100g/min) was significantly associated with being 61% more likely to exhibit fatigability independent of age and sex (Table 2). The odds ratio (OR) of CMRO_2_ remained significant after adjusting for ATPmax, VO_2_peak and weight (Table 2). Results were similar after adjusting for medication count and multimorbidity, albeit the association was no longer significant, p<0.05. OR for CMRO_2_ only minimally changed after adjusting for either max OXPHOS or max ETS (from 1.61 to 1.51 and 1.50, respectively, Table 2). Further adjustment for VO_2_peak, weight, medication count, and multimorbidity attenuated the results, p>0.05 (Table 2).

**Table 2:** Logistic Regression of the Association between Cerebral Metabolic Rate of Oxygen (CMRO_2_) and Accelerometry-Based Fatigability Status using the Pittsburgh Fatigability Index (PPFI) from the Fast-Paced 400m Walk: Study of Muscle, Mobility and Aging (SOMMA) – Brain Ancillary Study.

| **Models** | **Odds Ratio\*\***<br>**per 1 SD**<br>**CMRO<sub>2</sub>\*** | **95%**<br>**Confidence**<br>**Interval** | **P-**<br>**value** |
| --- | --- | --- | --- |
| CMRO<sub>2</sub> + age + sex (n=117) | 1.61 | (1.06, 2.45) | 0.03 |
| CMRO<sub>2</sub> + age + sex + ATPmax (n=105) | 2.00 | (1.23, 3.24) | 0.005 |
| CMRO<sub>2</sub> + age + sex + ATPmax + VO<sub>2</sub>peak (n=104) | 1.88 | (1.10, 3.21) | 0.02 |
| CMRO<sub>2</sub> + age + sex + ATPmax + VO<sub>2</sub>peak + weight (n=104) | 1.85 | (1.07, 3.20) | 0.03 |
| CMRO<sub>2</sub> + age + sex + ATPmax + VO<sub>2</sub>peak + weight +<br>medications + multimorbidity (n=103) | 1.75 | (0.99, 3.08) | 0.056 |
| CMRO<sub>2</sub> + age + sex + max OXPHOS + technician (n=105) | 1.52 | (0.98, 2.34) | 0.06 |
| CMRO<sub>2</sub> + age + sex + max OXPHOS + technician +<br>VO<sub>2</sub>peak (n=104) | 1.32 | (0.80, 2.17) | 0.3 |
| CMRO<sub>2</sub> + age + sex + max OXPHOS + technician +<br>VO<sub>2</sub>peak + weight (n=104) | 1.28 | (0.76, 2.15) | 0.4 |
| CMRO<sub>2</sub> + age + sex + max OXPHOS + technician +<br>VO<sub>2</sub>peak + weight + medications + multimorbidity (n=103) | 1.21 | (0.71, 2.07) | 0.5 |
| CMRO<sub>2</sub> + age + sex + max ETS + technician (n=105) | 1.50 | (0.97, 2.31) | 0.07 |
| CMRO<sub>2</sub> + age + sex + max ETS + technician + VO<sub>2</sub>peak<br>(n=104) | 1.32 | (0.80, 2.18) | 0.3 |
| CMRO<sub>2</sub> + age + sex + max ETS + technician + VO<sub>2</sub>peak +<br>weight (n=104) | 1.28 | (0.76, 2.16) | 0.3 |
| CMRO<sub>2</sub> + age + sex + max ETS + technician + VO<sub>2</sub>peak +<br>weight + medications + multimorbidity (n=103) | 1.22 | (0.71, 2.09) | 0.5 |

\*1 SD CMRO<sub>2</sub> = 2.87 µmol/100 g/min
\*\*Odds ratio of exhibiting fatigability (PPFI >0) vs no fatigability (PPFI=0, reference)
*Note: In addition to CMRO<sub>2</sub>, ATPmax, max OXPHOS, max ETS and VO<sub>2</sub>peak were entered into models as standardized variables. For models adjusted for weight, units of VO<sub>2</sub>peak are mL/min, in all other models, units are mL/kg/min.*

## Discussion

Our results suggest higher CMRO_2_ may influence performance fatigability in this cohort of community-dwelling older adults, independent of age and sex, muscle energetics and CRF. There is an ongoing debate as to whether higher oxygen consumption, irrespective of the tissue or organ being examined, may be compensatory or reflect de-differentiation. Higher oxygen extraction is associated with higher neural activation, and this in turn has been related to less mental fatigue (Darnai et al. 2023). Conversely, higher CMRO_2_ in older age may reflect reduced cellular metabolic efficiency (Peng et al. 2014; Lu et al. 2011). Our results are consistent with a study in patients with multiple sclerosis, showing higher CMRO_2_ in relation to fatigue (West et al. 2020); this positive association was interpreted as a response to metabolically active inflammatory processes demanding greater oxygen extraction, and causing worse symptoms and greater fatigue. CMRO_2_ may also increase in response to impairments occurring systemically (e.g., at cardiopulmonary level). In this sample, higher CMRO_2_ was significantly correlated with lower VO_2_peak, and this in turn was associated with higher fatigability. Future studies should assess whether higher CMRO_2_ may be a central response to lower CRF.

Given the role of muscle energetics and CRF on fatigability, we assessed to what extent the relation between CMRO_2_ and PPFI was modified when accounting for these variables. Although the main association remained statistically significant, we also found the influence of muscle energetics varied depending on the metric used. The association between higher CMRO_2_ and fatigability was strengthened (22% higher) when ATPmax was added to the model, whereas it was only minimally attenuated after the addition of ex vivo muscle energetics measures. This may be because ATPmax is an in vivo measure of maximal ATP production during muscle contraction (Coen et al. 2013), whereas respirometry metrics assess mitochondrial function in ideal conditions. Thus, ex vivo measures may not be strong predictors of performance when objective measures of brain energetics, fatigability and CRF are in the models. Another possibility is due to statistical power.

Participants that exhibited fatigability were 7 kg heavier than those with no fatigability. After adjusting for weight, the relation between CMRO_2_ and fatigability was attenuated. Prior work corroborates our results by revealing an association between higher body mass index and greater perceived physical fatigability in a nationally representative sample of adults aged 60-64 (Cooper et al. 2019). Those exhibiting fatigability took more medications and had more medical conditions than those without fatigability, and this may explain why adjustment for these variables attenuated the association of CRMO_2_ with PPFI.

The SOMMA-Brain cohort is healthier than the general population (Rosano et al. 2024) which may limit generalizability of our work. Note that even in our healthier than average cohort CMRO_2_ was significantly associated with fatigability after adjusting for in vivo muscle energetics and CRF. Thus, in a less healthy population, the magnitude of the association may be larger than in our findings. Limitations of our work include our cross-sectional analysis; thus, we were unable to determine temporality between CMRO_2_ and fatigability. Future work should evaluate whether changes in CMRO_2_ predict changes in fatigability in older adults, or vice versa. Further, CMRO_2_ was quantified at the whole-brain level. Thus, metabolic changes in specific regions of the brain may be driving the observed association. While other measures provide regional energy metabolic indices, they present numerous limitations for studies in older adults (Paling et al. 2011). Proton spectroscopy has limited specificity to energy loss, because the metabolites are also linked to parenchymal damage; and perfusion methods require port access which is high risk, especially for older adults. Compared to other methods, our neuroimaging protocol is non-invasive and requires relatively short scan times (<15 minutes). Higher tolerability of the protocol reduces nonparticipation and bias and increases the probability of capturing a more representative sample.

Our work is strengthened by the use of accelerometry-based fatigability. PPFI applies weights to emphasize performance decrement at the beginning of the walk and to limit motivation effects at the end of the walk. PPFI considers the entire trajectory of the walk when determining maximum cadence and calculating performance fatigability. We also used both in vivo and ex vivo measures of muscle energetics and gold-standard fitness testing.

Emerging studies show brain energy metabolism can respond to behavioral and nutritional interventions in older adults (Zhou et al. 2018; Haeger et al. 2020; Matura et al. 2017; Balestrino & Adriano 2019; Tardy et al. 2020), but their effect on fatigability have not been investigated. Future research should focus on longitudinal associations and identification of metabolic changes in specific regions of the brain, as it is known that metabolic changes in the prefrontal cortex are associated with fatigue and cognitive function in those with multiple sclerosis (Zuppichini et al. 2023). Additionally, we found that higher perceived physical fatigability was associated with smaller hippocampal, putamen, and thalamus volumes (Wasson et al. 2019). Thus, evaluating regional measures of CMRO_2_ in these locations may identify the metabolic changes behind the observed relation between brain energetics and fatigability in older adults. Our findings indicate that a multi-symptom approach is needed to better understand the biology of performance fatigability in older adults.

## Data Availability

All data produced in the present study are available upon reasonable request to the authors.

https://sommaonline.ucsf.edu/

## Author Contributions

Ms. Gay and Drs. Rosano and Glynn had full access to all of the data for the study and take responsibility for the integrity of the data and accuracy of the data analyses. All authors: interpretation of data, critical revision of manuscript for important intellectual content. All authors read and approved the submitted manuscript.

## Data Availability Statement

SOMMA data are publicly available by request at https://sommaonline.ucsf.edu/.

